# Merging the Traditional with the New *CME-accredited Twitter Journal Clubs*

**DOI:** 10.1101/2020.04.22.20075606

**Authors:** Tejas Desai

## Abstract

Twitter journal clubs are increasingly popular amongst healthcare providers. Most journal clubs rely on voluntary physician participation. Offering continuing medical education credit may incentivize and improve these journal clubs.

In this investigation a series of 5 consecutive publication-sponsored Twitter journal clubs were analyzed in calendar year 2016, in which the latter 3 journal clubs offered CME credit. Various quality metrics were measured and analyzed to identify sustainable improvements in those journal clubs that offered CME credit.

Overall, Twitter journal clubs that offered CME credit performed better in certain quality metrics, to wit activity, originality, and evidence-based tweeting, but fared poorly in number of and type of participant interactions.

Twitter journal clubs are in their infancy and physician participation remains steady. Offering CME credit improves certain quality metrics within these journal clubs. This investigation should encourage more publications to sponsor CME-accredited Twitter journal clubs.

## INTRODUCTION

The medical journal club (JC) remains an important educational exercise for healthcare providers (HCP). Journal clubs allows individuals with a variety of expertise to discuss, debate, and contemplate the clinical and scientific impact of medical science [1]. Indeed from its inception over a century ago, the JC can be considered the first iteration of social media in medicine. Prior to its adoption as an educational tool, HCPs absorbed medical information in near isolation. The JC broke this solitary learning confinement by physically bringing together providers to share their perspectives, interpretations, and experiences with one another. So valuable is the JC that for the better part of half-acentury, the format and manner in which it has been conducted has remained rather unchanged [2].

In the last one to two decades the JC has evolved. By incorporating audio and video teleconferencing, a larger number of HCPs are capable of participating in their local JC. More recently, the notion that JC participants need to be geographically and temporally local to the JC host has been challenged by the implementation of Twitter JCs [2-4].

Having all the traditional hallmarks of the well-familiar medical JC (i.e., an article to discuss, pre-specified topics, pre-selected moderator(s), well-defined duration), the Twitter JC operates at a higher level of recruitment. HCPs from around the globe participate and as a treat, the article author(s) join the Twitter JC to enhance discussion and offer unpublished insights [1,2,5,6]. Skeptics should note that the success of the Twitter JC is no more readily apparent than in the increasing number and frequency of medical disciplines adopting and hosting the Twitter JC in the past two years [1,7].

Consider that a select few faculty commonly host the medical JC and one quickly recognizes the egalitarian nature of the Twitter JC [8,9]. Non-academically affiliated individuals, who were traditionally viewed as JC consumers, band together to manage, promote, host, and moderate regularly scheduled Twitter JCs [1,2]. The anecdotal juxtaposition with traditional JCs is eye opening: Twitter JCs reach a global audience two times greater or more than the local JC [2,7,10,11]. Perhaps this contrast alone has convinced a number of medical journals to conduct their own Twitter JCs. Publication-sponsored Twitter JCs are a product of and a contributor to the cumulative success of all Twitter JCs [9,12-15].

Most recently, an ambitious and uniquely novel feature of the Twitter JC has emerged. Since its inception Twitter JC participants have been rewarded through the following intangible benefits: knowledge, community inclusion, and networking opportunities. In calendar year 2016, the Journal of Hospital Medicine (@JHospMedicine) became the first Internal Medicine, PubMed-indexed journal to reward participants with Category 1 AMA continuous medical education (CME) credit for participation in their Twitter JCs. This investigation hypothesizes that the participation and interaction in CME-accredited Twitter JCs will be far greater than in a standard Twitter JCs [2,16].

## METHODS

This investigation analyzes data from five Twitter JCs hosted by the Journal of Hospital Medicine (Journal) in calendar year 2016. The Journal hosted its first Twitter JC on 12-October-2015. Data from this JC was excluded from analysis because it occurred in a calendar year different than the remaining five. The CME- unaccredited Twitter JCs occurred on 11-January and 4-April, while the CME-accredited Twitter JCs occurred on 11-July, 12- September, and 7-November. The author collected and analyzed all publicly available tweets and their corresponding metadata containing the journal-club-specific hashtag #JHMChat during each JC session. Although each session was scheduled for one hour in duration, frequently the sessions started and/or ended outside of this 60-minute window. As a result, this author analyzed the content of each tweet to determine the start and end of each Twitter JC (Table 1). Chronologic ordering of tweets resulted in five subsets containing tweets that were authored between the start and end tweets; this investigation analyzes those subsets. Identifying JC moderators occurred by analyzing: 1) the author of and accounts mentioned in the start tweet, 2) the accounts used to announce topics, and 3) the author of the end tweet. Pursuant to this method, more than one moderator account was identified in some Twitter JCs. Identifying article authors and expert discussants occurred by analyzing the tweet content of the moderator(s) for the author/discussant’s Twitter account name.

**Table 1:**
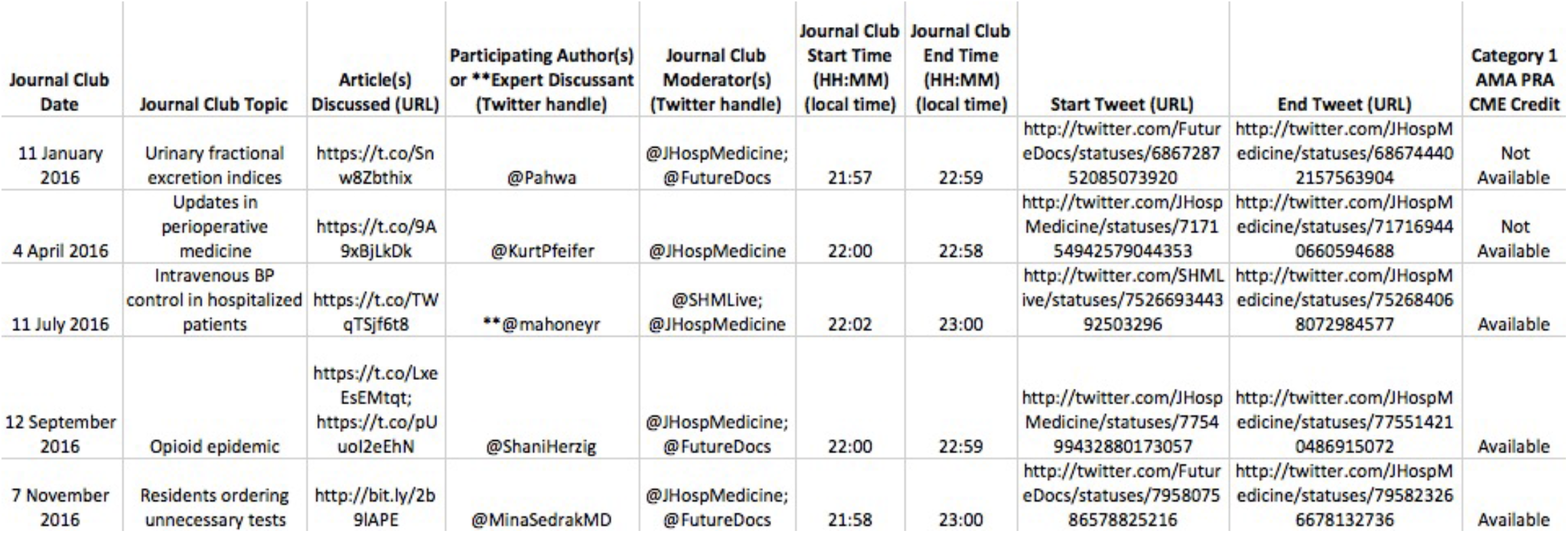
Journal Clubs Analyzed.

This author hypothesized that there would be a sustained improvement in the participation, originality, scientific support, and/or interactions within the Twitter JC once CME-credit was offered. The following quality metrics were analyzed to test this hypothesis: 1) number of participants, 2) number and ratio of original tweets and retweets, 3) number of evidence-based tweets, and 4) number and type of participant-participant interactions [2,14,17,18]. This author defined a participant as a Twitter user/account that authored at least one tweet during the JC. A sustained increase in the number of participants would support the hypothesis. Second, this investigation employed the commonly accepted definitions of an original tweet and retweet: an original tweet was a unique and new tweet authored by a participant, while a retweet was a copied and rebroadcasted tweet by a different participant. Based on work by Widmer et al, a higher original tweet to retweet ratio would indicate a larger amount of new and unique content in the JC [12]. Third, the author employed and expanded upon Djuricich’s definition of an evidence-based tweet to include tweets with either hyperlink citations to or an attached figure or table from the medical literature [2,18-20]. This author, as well as other investigators, considered the quality of the Twitter JC to be directly related to the amount of evidence-based tweets. Finally, any participant who directed his/her tweet to another participant created a relationship with the second participant. This author counted each instance of such a relationship as a single participant-participant interaction. A larger number of interactions would support the hypothesis.

This author intentionally avoided performing statistical analyses of the data. Given the small number of Twitter JCs that offer CME-credit and the small number of tweets to analyze within each one-hour JC, the author believed that any statistically significant finding would be un-interpretable at best and misleading at worst. The author attempted to increase the sample size to allow for meaningful statistical analyses, but the uniqueness of the JHM-hosted Twitter JCs hampered this effort. To the best of this author’s knowledge, no other PubMed-indexed publication- or independently-sponsored Twitter JC offers CME credit.

Finally, this author discloses that he neither had nor holds any relationship with the Journal of Hospital Medicine. In an attempt to avoid any appearance of bias, this author has not submitted any prior iteration of this investigation to the Journal or its representatives for consideration.

## RESULTS

There were five Twitter JCs analyzed: the first two did not offer CME-credit. A total of 1,619 tweets were analyzed from 193 participants during calendar year 2016. Less than half of all tweets were composed during the non-CME-accredited journal clubs (607 tweets; 37%). A similar proportion of individuals participated in the non-CME-accredited journal clubs (38%). The 11-July journal club, which was the first journal club to offer CME-credit, contained the largest number of tweets (374 tweets; 23%). Moreover, this journal club was the only one of the five studied in which an expert discussant participated in lieu of the article authors (Table 2).

**Table 2:** Journal Club Metrics.

| Chat Date | Total Tweets Composed (Number) | Participants (Number) | Total Tweets Composed by Moderator(s) (Number) | Total Tweets Composed by Moderator(s) (Percent of Total Tweets) | Tweets Composed by Article Author or Expert Discussant (Number) | Tweets Composed by Article Author or Expert Discussant (Percent of Total Tweets) | Original Tweets (Number) | Retweets (Number) | Original Tweet:Retweet Ratio | Total Tweets with Figures or Tables (Number) | Total Tweets with hyperlinked citations (Number) | Total Evidence-Based Tweets (Number) | Total Evidence-Based Tweets (Percent of Total Tweets) | Participants Involved in a direct Participant-Participant Interaction (Number) | All Participant-Participant Interactions (Number) | Specific Participant-Moderator Interactions (Number) | Specific Participant-Author or Expert Discussant Interactions (Number) |
| --- | --- | --- | --- | --- | --- | --- | --- | --- | --- | --- | --- | --- | --- | --- | --- | --- | --- |
| 11 January 2016 | 331 | 53 | 79 | 23.87 | 37 | 11.18 | 176 | 155 | 1.14 | 8 | 70 | 78 | 23.56 | 40 | 435 | 100 | 70 |
| 4 April 2016 | 276 | 21 | 90 | 32.61 | 39 | 14.13 | 163 | 113 | 1.44 | 11 | 42 | 53 | 19.20 | 23 | 292 | 52 | 76 |
| 11 July 2016 | 374 | 57 | 93 | 24.87 | 26 | 6.95 | 218 | 156 | 1.40 | 55 | 106 | 161 | 43.05 | 42 | 412 | 91 | 68 |
| 12 September 2016 | 275 | 31 | 101 | 36.73 | 35 | 12.73 | 169 | 106 | 1.59 | 8 | 99 | 107 | 38.91 | 29 | 273 | 57 | 75 |
| 7 November 2016 | 363 | 31 | 87 | 23.97 | 20 | 5.51 | 271 | 92 | 2.95 | 7 | 73 | 80 | 22.04 | 38 | 404 | 82 | 48 |

The 11-July journal club had the largest number of participants at 57, followed by 51 participants in the 11-January JC. The last CME-accredited journal club of the year, 7-November, had 3% fewer tweets and nearly 45% fewer participants than the 11-July journal club. The last non-CME-accredited journal club of the year, 4-April, had 17% fewer tweets and 60% fewer participants than the 11-January JC. The greatest activity from the general audience was seen in two of three CME-accredited journal clubs, as evident by the low percentage of tweets composed by the moderator(s) and author(s)/expert discussant. In the 11-July and 7-November JCs, moderator(s) activity was less than 25% and author(s)/expert discussant activity was less than 7% of total tweets composed (Table 2).

An often-undesirable quality of Twitter journal clubs is to have more retweets than original tweets. All of the JCs in this investigation had more original than repetitious content. Two of the top three JCs with the greatest proportion of original tweets were CME-accredited (7-November: 2.95 OT:RT; 12-September: 1.59 OT:RT). The 11-January JC was the least original journal club within the data set (1.14 OT:RT). Interestingly, the 7-November JC, which had the greatest proportion of original content (highest OT:RT ratio) also had the second lowest proportion of evidencebased tweets (22%). Nearly half of all tweets were supported by evidence in the 11-July JC; the largest percentage within the data set. There was downward trend in scientific support in subsequent CME-accredited journal clubs (Table 2).

In two Twitter JCs there were a greater number of participants involved in at least one interaction than the total number of participants involved in the JC itself (4-April and 7-November). The difference between these two values represents the number of individuals who were “asked” or “recruited” (directly or indirectly) to join the Twitter JC but chose not participate (4-April: 2 individuals; 7-November: 7 individuals). While nearly all of the participants in the 12-September JC were involved in a participant-participant interaction, CME-unaccredited JCs had the greatest number of interactions (Figure 1). The 11-January JC had nearly 6% more total interactions and 10% more participant-moderator (PM) interactions than the 11-July JC (Total: 435 versus 412, respectively; PM: 100 versus 91, respectively) (Figure 2). The number of interactions between participants and the article author or expert discussant was greatest in the 4-April JC.

**Figure 1:**
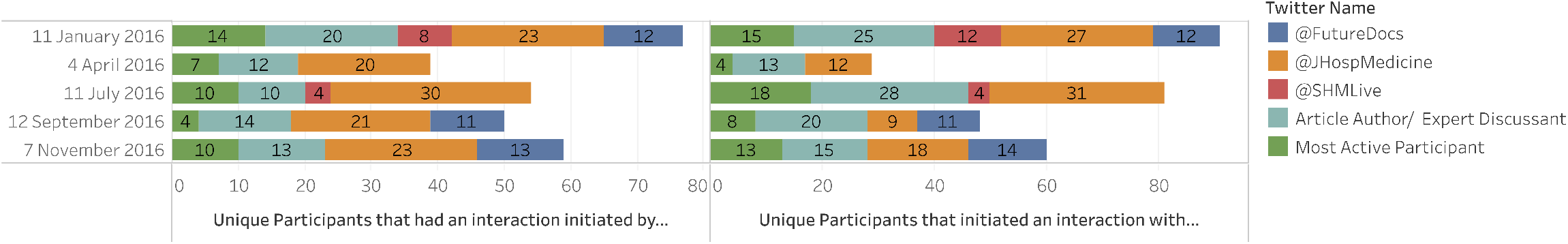
Types of Journal Club Interactions.

**Figure 2:**
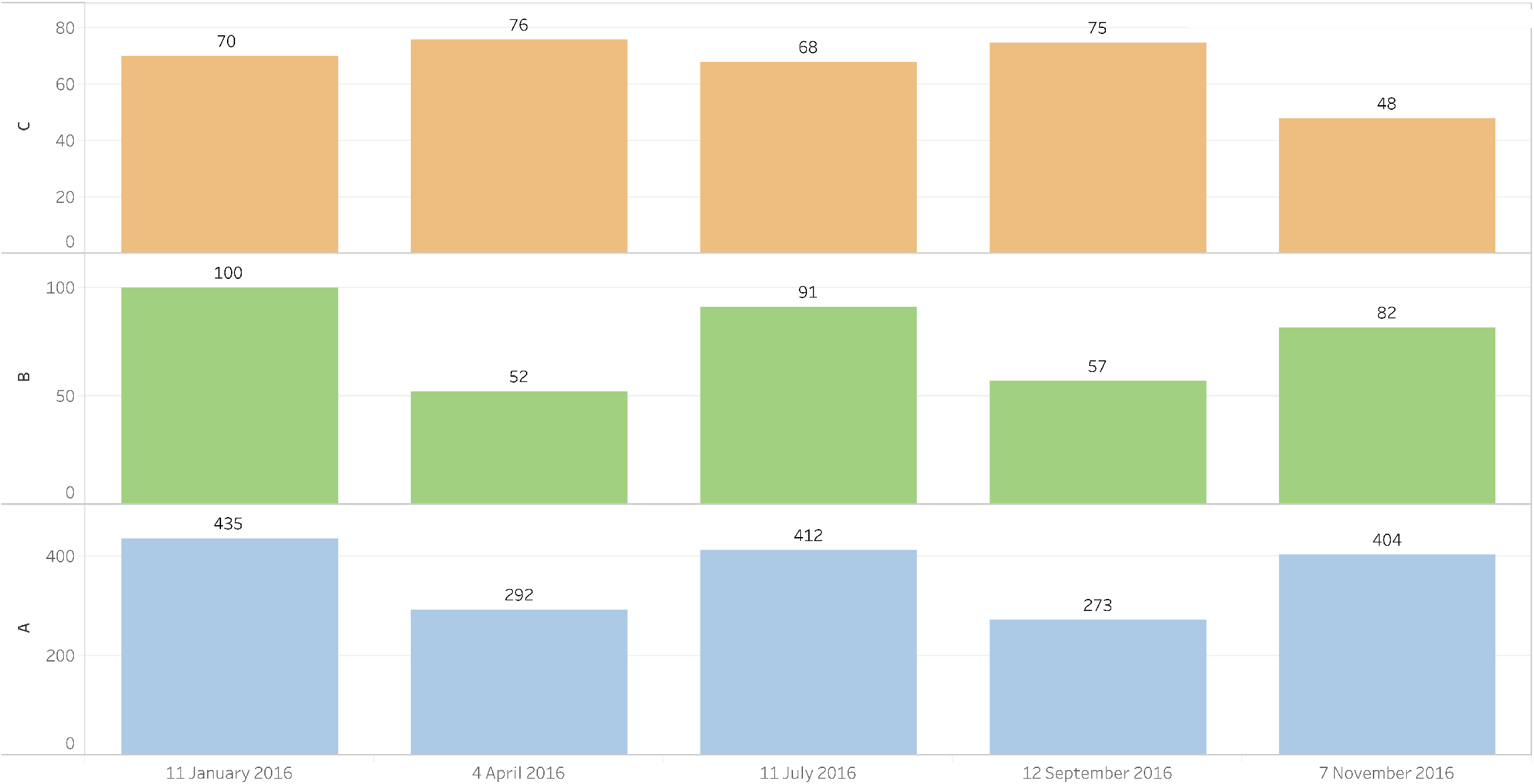
Number of Journal Club Interactions.

## DISCUSSION

Twitter journal clubs in which continuing medical education credit was available generally outperformed those journal clubs that did not offer the credit. However, there does not appear to be a trend toward consistently and increasingly outperforming CME-unaccredited journal clubs. In the eight metrics of quality shown in Table 3, the CME-accredited Twitter JCs ranked highest in five and second highest in six. Specifically, CME-accredited JCs did better with tweet activity, originality, and evidence-based tweeting: ranking first in these metrics. Conversely, CME-unaccredited JCs outperformed in the number and type of interactions (Figures 3-7). The unaccredited JCs had more interactions between participants and both moderator(s) and article author(s) or expert discussant than the CME-accredited JCs.

**Table 3:** Quality Metrics and Journal Club Ranking (shaded chats are CME-accredited)

|  | Ranks 1st | Ranks 2nd | Ranks 3rd |
| --- | --- | --- | --- |
| **Total Tweets** | 11 July | 7 November | 11 January |
| **Total Number of Participants** | 11 July | 11 January | 12 September & 7 November |
| **OT:RT Ratio** | 7 November | 12 September | 4 April |
| **Percent Evidence-Based Tweets** | 11 July | 12 September | 11 January |
| **Number of Participants Involved in an Interaction** | 11 July | 11 January | 7 November |
| **Total Participant-Participant Interactions** | 11 January | 11 July | 7 November |
| **Total Participant-Moderator Interactions** | 11 January | 11 July | 7 November |
| **Total Participant-Author/Expert Discussant Interactions** | 4 April | 12 September | 11 January |

**Figure 3:**
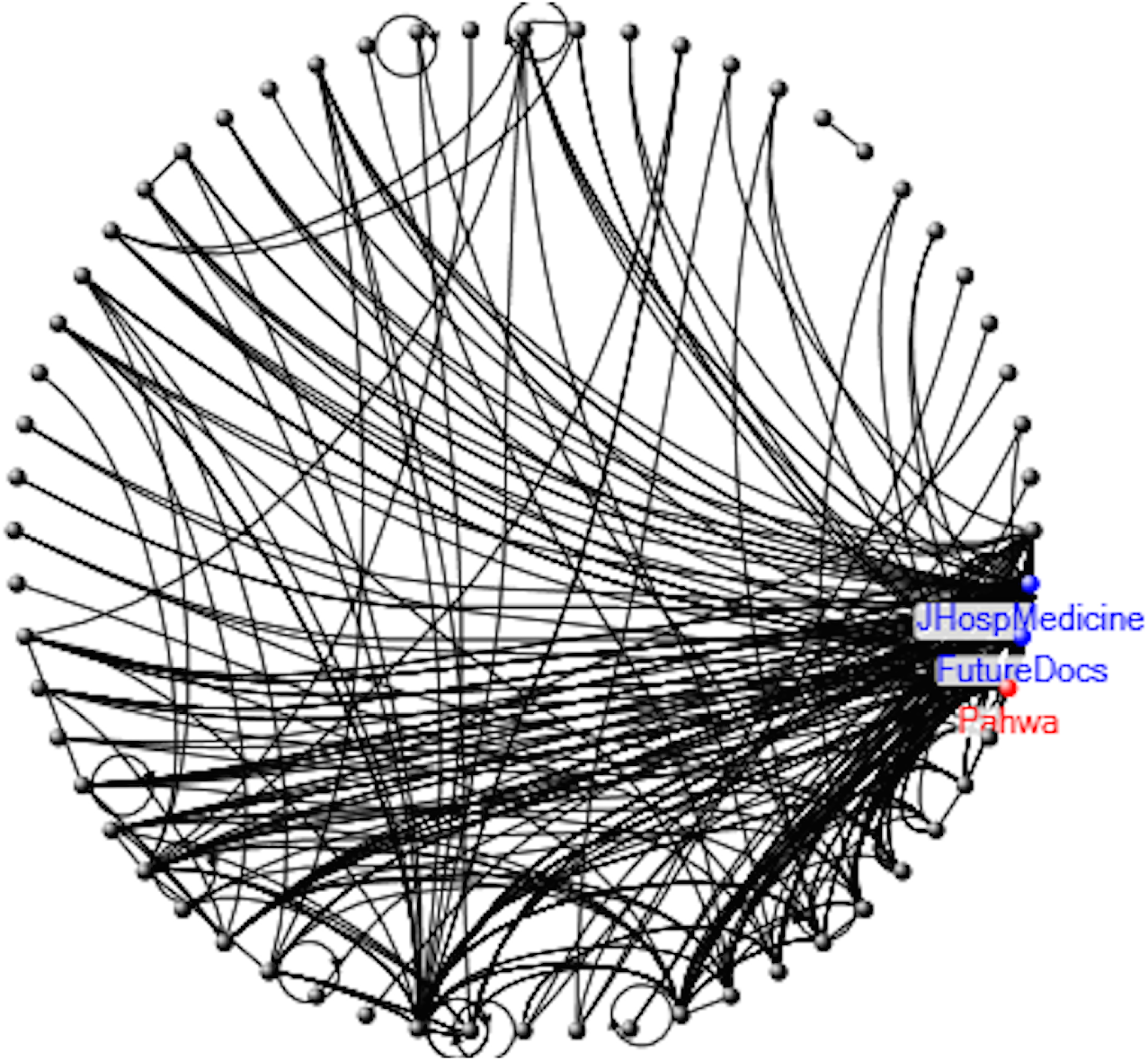
Interaction Map of the 11 January 2016 Twitter Journal Club.

**Figure 4:**
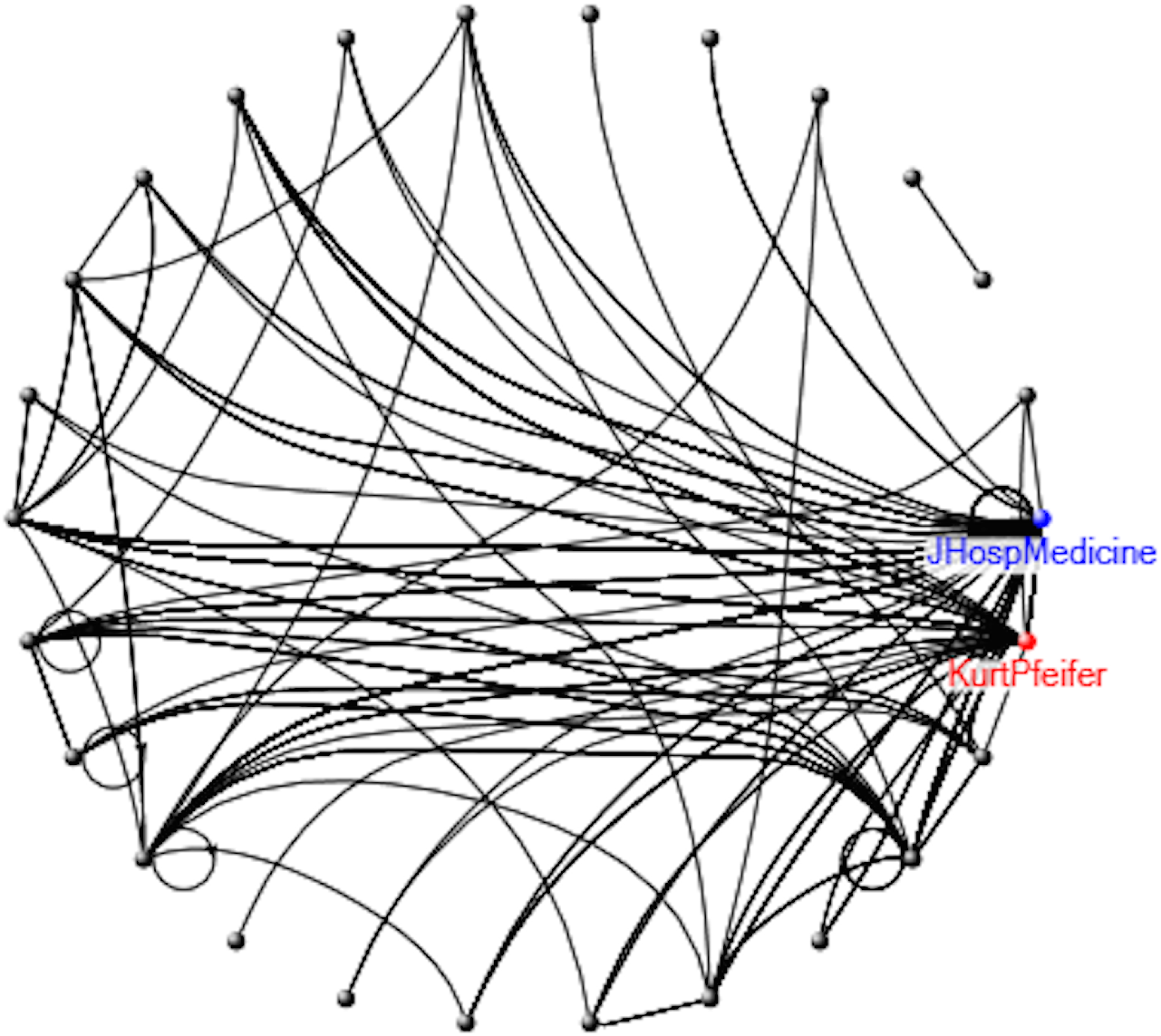
Interaction Map of the 4 April 2016 Twitter Journal Club.

**Figure 5:**
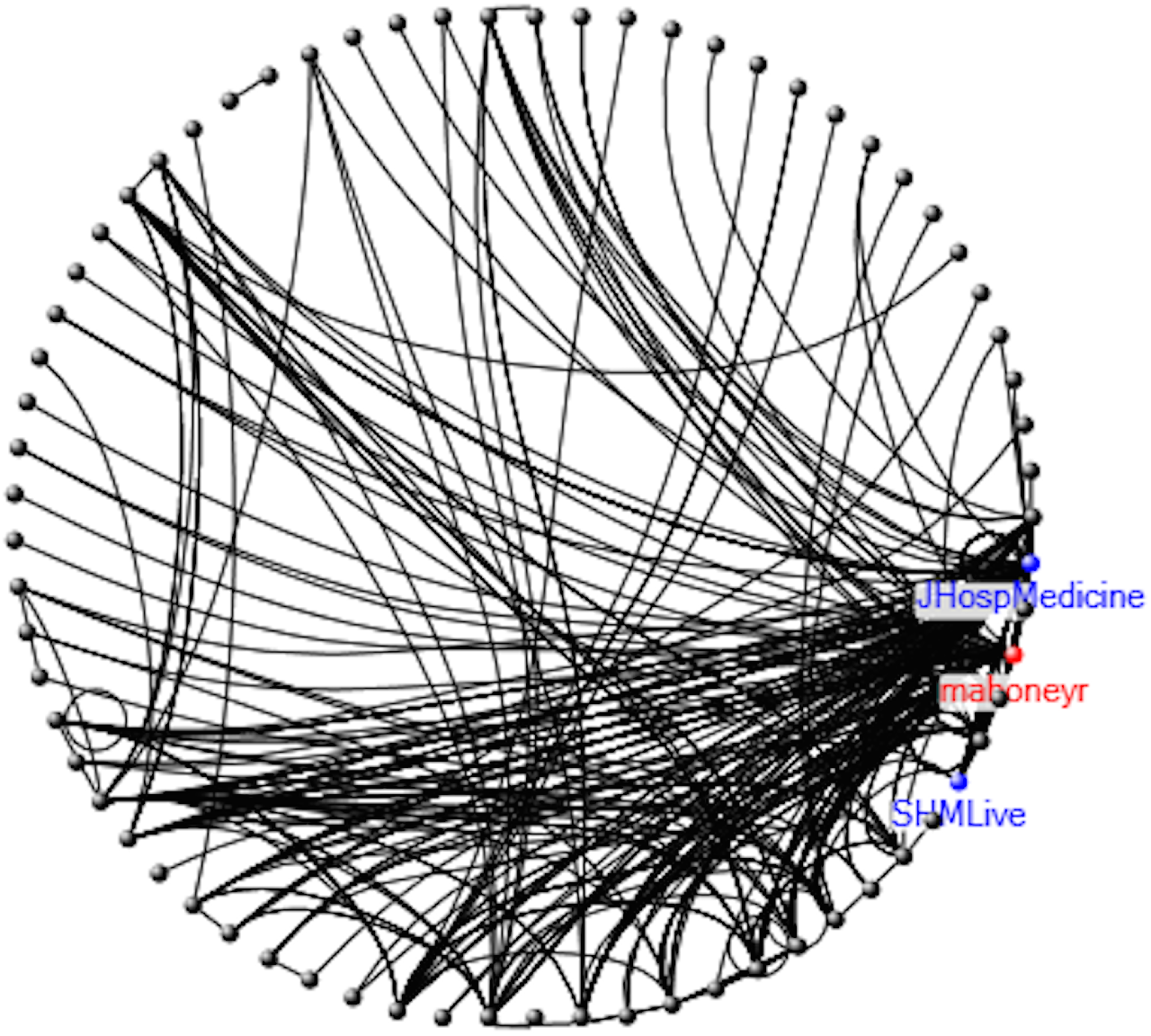
Interaction Map of the 11 July 2016 Twitter Journal Club.

**Figure 6:**
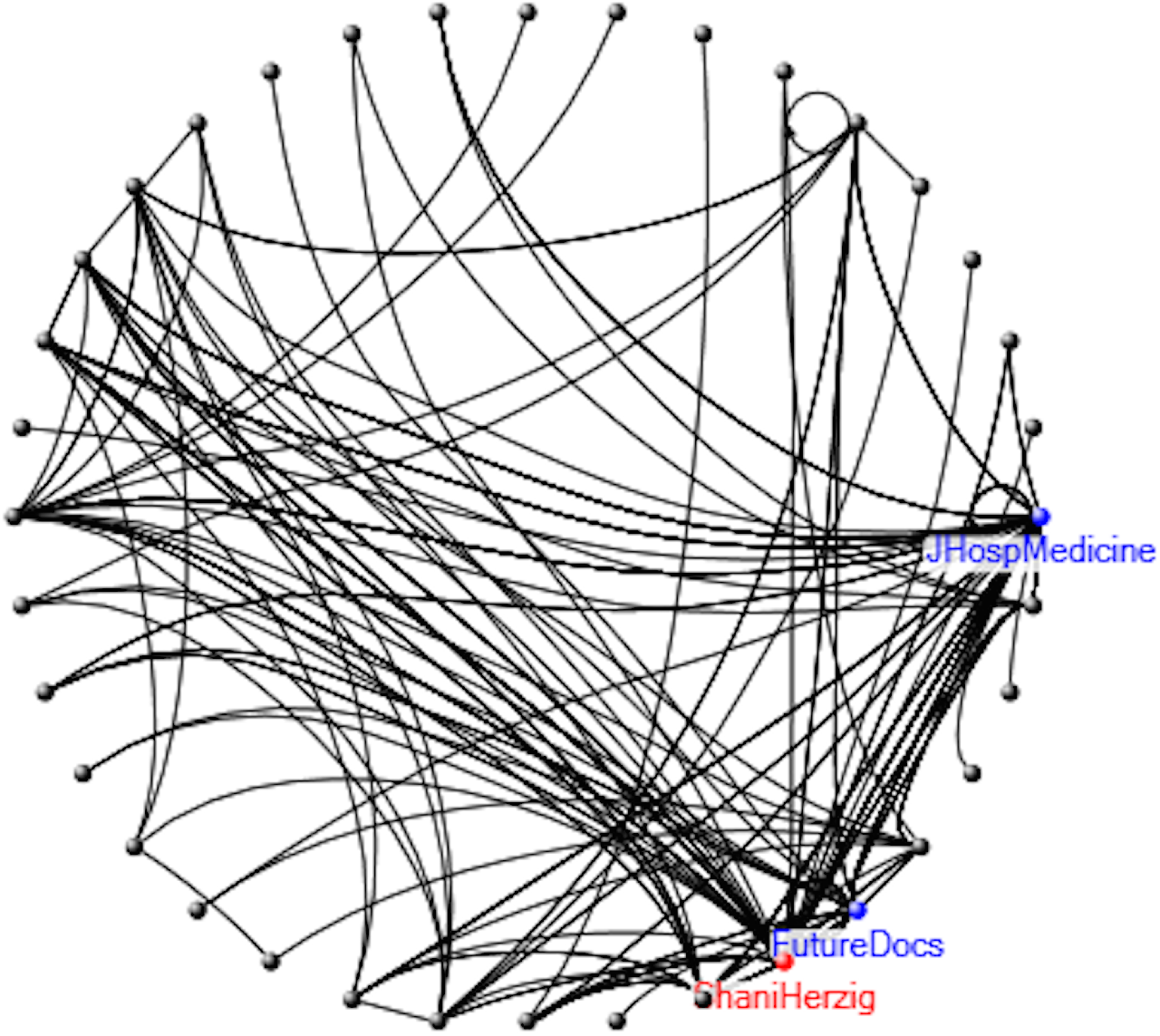
Interaction Map of the 12 September 2016 Twitter Journal Club.

**Figure 7:**
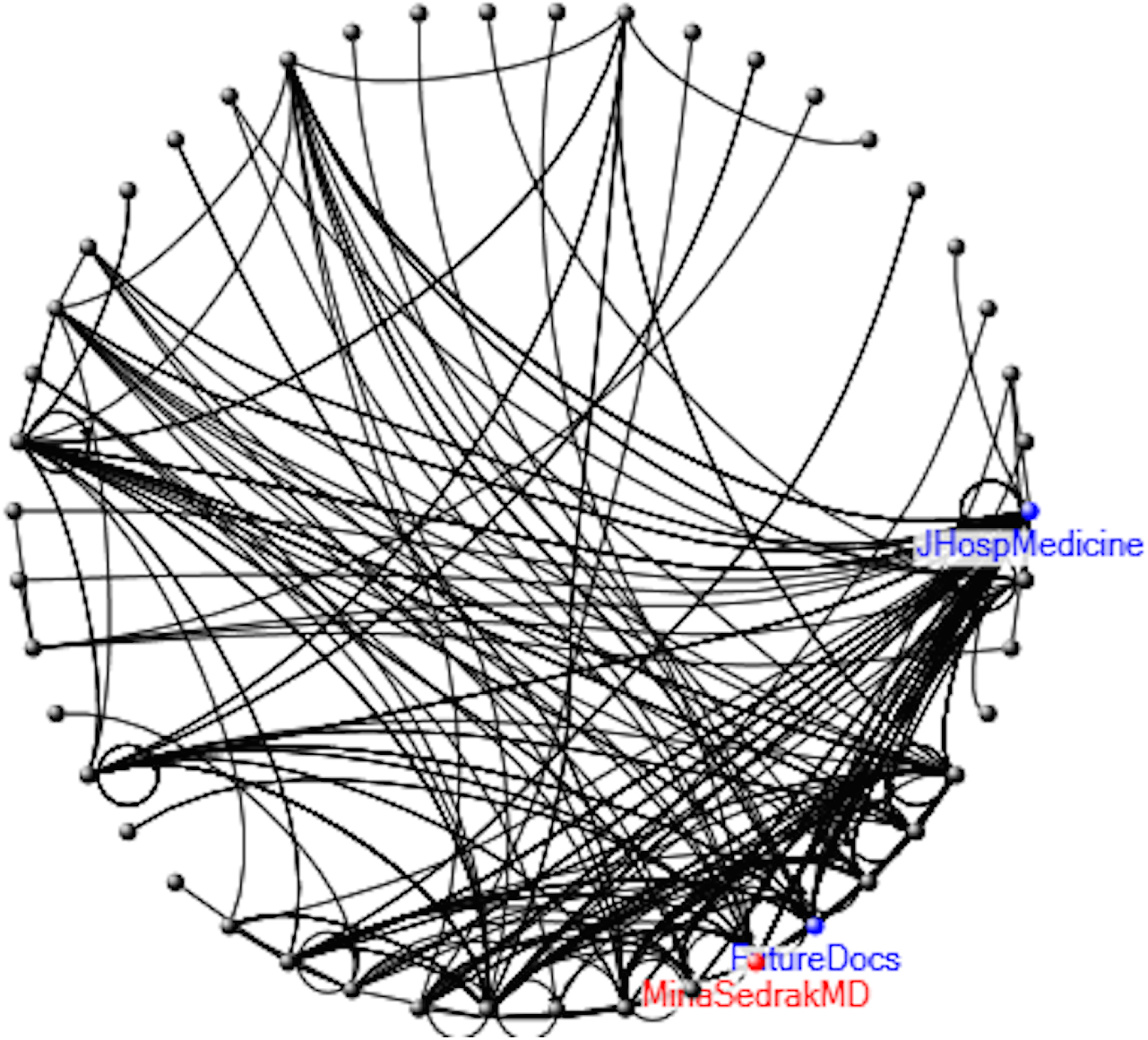
Interaction Map of the 7 November 2016 Twitter Journal Club.

The inconsistency in outperforming CME-unaccredited Twitter journal clubs may lead some readers to think that the availability of CME credit does not add value to such journal clubs. This belief would be contrary to what Chen and others have suggested: that offering some incentive for participation would improve Twitter journal clubs [2]. The data in this investigation show that CME availability did improve some areas of quality but had a much less positive effect on interactivity. In an attempt to resolve this dichotomy, this author raises two alternative explanations for the findings in this report. First, it is reasonable to view CME availability less as an incentive and more as recognition for physician participation. The process in which one reads, synthesizes, and responds to a rapid stream of scientific tweets (some that include external references and/or figures/tables) is admirable, educationally valuable, and worthy of tangible recognition (CME credit). Second, it is possible that another mechanism, unrelated to CME availability, exists by which sponsors can improve Twitter JCs. The data in this investigation raise the possibility that having an expert discussant, rather than the article author(s), participate in the JC can lead to a higher quality JC. The 11-July JC was the only JC in the examined data set that replaced article author(s) involvement with an expert discussant. When compared to the remaining two CME-accredited JCs, the 11-July JC scored the highest in four of the five first-place rankings and six of eleven first-and-second-place rankings. Indeed Chen and others have hypothesized that the presence of article authors can “stifle” interaction between JC participants for fear of insulting or negatively critiquing the authors’ work [2] (Figure 8). The data presented here offers the first quantified support of this hypothesis.

**Figure 8:**
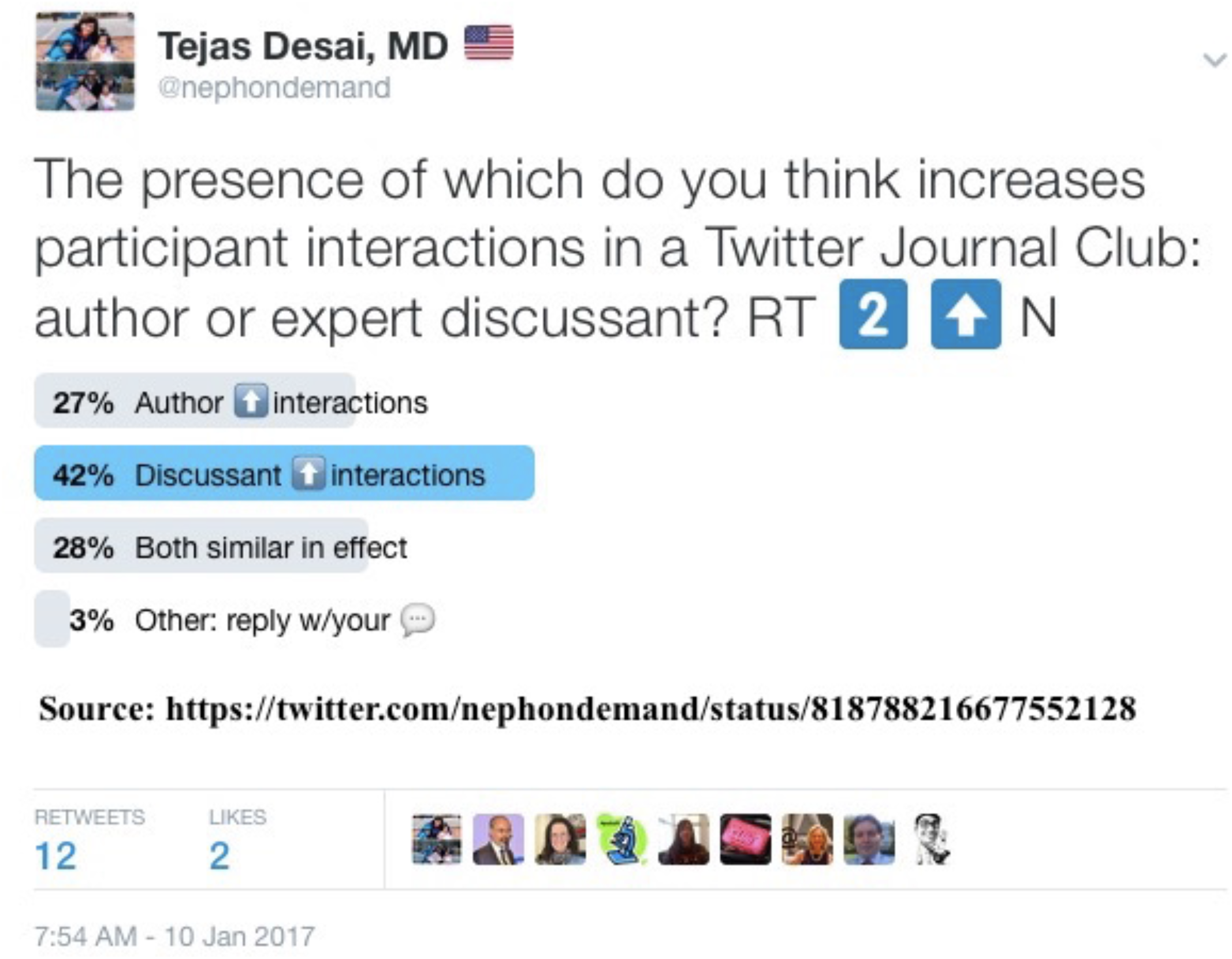
Twitter Poll.

While there are more independently-sponsored than publication-sponsored Twitter JCs, the Journal of Hospital Medicine is not the only Internal Medicine-focused publication that conducts regular Twitter JCs. Indeed the Journal of General Internal Medicine (JGIM) and Journal of Graduate Medical Education (JGME) have and continue to host Twitter JCs. The extent of data reporting by these respective Journals is sparse and that creates difficulty in evaluating various interventions intended to improve JC quality [9,14]. Mehta and Flickinger reported the following data from the first JGIM Twitter JC on 5 June 2014: 367 tweets from 78 participants in 60 minutes [9]. Thereafter additional data, if available, has not been publicly released. Lin and Sherbino reported on the first JGME sponsored Twitter JC in January 2015 and admitted that “web traffic” was the best way to quantify JC success [14]. Publications in other medical specialties have sponsored Twitter JCs with varying frequency but none have reported extensively on quality metrics as detailed herein.

## CONCLUSION

Offering CME credit improved specific quality metrics in the Twitter journal clubs analyzed. Given these improvements, more publications should consider sponsoring CME-accredited Twitter JCs. Indeed at a minimum CME credit should continue to be offered to reward physicians who participate in these online activities.

## Data Availability

Data available upon request

## ARTICLE INTERACTIVITY

Below are the types of interactions generated by this investigation. Tweet prefix is **https://twitter.com/nephondemand/status/**

### Retweets of */827928021243944965*

- Tulun Sokit @tulunsokit
- rheuMD @reinmd77
- Hector Madariaga, MD @HecmagsMD
- Basu Gopal @BasuNephro
- Holly Caretta-Weyer @holly_cw
- Teresa Chan @TChanMD
- Sivakumar Sridharan @sayitmyway
- Arvind Canchi @arvindcanchi
- Szymon Brzósko @brzoskos
- Edoardo Melilli @EdoardoMelilli
- Errant Nephron @Errantnephron
- linaAl @lina_alimari

### Retweets of */828236235365814272*

- Scherly Leon, MD @SLeonMD
- Kaitlin Sundling @KaitlinSundling
- Ultimate Librarian @ultimatelibrarn
- Mike Thompson, MDPhD @mtmdphd
- Nancy H Stewart @nvhs0401
- Rolando Claure @RClaure_nefro
- Krishna Penmatsa @krishnadoctor1
- Tulun Sokit @tulunsokit
- Szymon Brzósko @brzoskos
- Cassie Rametta @CRcomment
- Kadrin Wilfong, MD @KadrinWilfong

### Retweets of */827932204395270144*

- Elaine Leung @elaineleung
- Basu Gopal @BasuNephro
- Doug @funnypop66
- Harvard Macy @harvardmacy
- Irsk Anderson @ijanders1
- Margaret Chisolm, MD @whole_patients
- Nora @noisy_nora
- Khalid Khan @Profkkhan
- Mike Thompson @mtmdphd

### Retweets of */828758549262110720*

- Michelle Brooks @michellebr00ks
- Errant Nephron @Errantnephron
- Fiona Loud @FionaCLoud

### Retweets of */831707620524306434*

- Alexandra Winter @alexandralw2

### Retweets of */847755646644744192*

- Ernesto Lopez Almaraz @kidney_doctor
- Audun Utengen @audvin

### Likes of */827928021243944965*

- Tulun Sokit @tulunsokit
- rheuMD @reinmd77
- Dr. Paul Sufka @psufka
- ananda reddy gurram @gurramanand
- Basu Gopal @BasuNephro
- Paul Raju @PaulRaju17
- Holly Caretta-Weyer @holly_cw
- Holden Caulfield PhD @ClinicalBriefs
- Sivakumar Sridharan @sayitmyway
- Fra Ian @caioqualunque
- Szymon Brzósko @brzoskos
- Edoardo Melilli @EdoardoMelilli
- Amit Langote @LangoteAmit
- Academic Life in EM @ALiEMteam

### Likes of */828236235365814272*

- Scherly Leon, MD @SLeonMD
- Soc. of Hospital Med @SHMLive
- george demetri @DrSarcoma
- Ultimate Librarian @ultimatelibrarn
- Dr. Deanna Attai @DrAttai
- Mike Thompson, MDPhD @mtmdphd
- Nancy H Stewart @nvhs0401
- Malvinder Parmar @wittykidney
- Errant Nephron @Errantnephron
- Szymon Brzósko @brzoskos
- Shoshana Herzig @ShaniHerzig
- Kenar Jhaveri @kdjhaveri
- Tony Breu @tony_breu
- Abdulmunim Aljapawe @2fbcb694e8164fd
- Bill Wood @WoodBD
- Cassie Rametta @CRcomment
- Susan Thomas @kidsbeansdoc

### Likes of */827932204395270144*

- Harvard Macy @harvardmacy
- Basu Gopal @BasuNephro
- Ryan Madanick, MD @RyanMadanickMD
- Szymon Brzósko @brzoskos
- Kenar Jhaveri @kdjhaveri
- Irsk Anderson @ijanders1
- Sambit Dash @sambit_dash
- Ali Jazayeri @majazayeri
- Margaret Chisolm, MD @whole_patients
- Mike Thompson @mtmdphd
- Arash Davari serej @arashdavari

### Likes of */828758549262110720*

- Michelle Brooks @michellebr00ks
- Nathaniel Reisinger @nephrothaniel
- Itunu Owoyemi @remyte7

### Likes of */831707620524306434*

- Dr. Cedrek McFadden @cedrekmd
- Heather Logghe, MD @LoggheMD
- Alexandra Winter @alexandralw2

## PEER REVIEW AND COMMENTARY

The team at NOD Analytics believes in reviewer-identified open-access peer review and commentary. To submit your review/comments, visit NOD Analytics or tweet @nephondemand. Submitted responses will be made immediately available to the reader in the space below.

- Jennifer Wyman @JenniferWyman4: #htech17 apropos our discussion about twitter journal clubs - quality and credit | https://twitter.com/JenniferWyman4/status/828286241049288705
- Robert Mahoney @mahoneyr: #JHMChat alumni make the prime time. Thanks for sharing! | https://twitter.com/mahoneyr/status/828241877967204354
- Red Hoffman, MD, ND @RedMDND: I just spent way too much time on your website- awe- some stuff! So glad to have discovered you! | https://twitter.com/RedMDND/status/831755299665092609
- Heather Logghe, MD @LoggheMD: Great resources on Twitter journal clubs! | https://twitter.com/LoggheMD/status/832323948675112960
- Amit Langote @LangoteAmit: Self publishing of research : The future of publications? #openaccess | https://twitter.com/LangoteAmit/status/834236380930781184
- Amit Langote @LangoteAmit: @nephondemand How can quality control be maintained? | https://twitter.com/LangoteAmit/status/834238182304710656 AUTHOR RESPONSE: Peer Review can be crowd-sourced & not anonymized. Identities matter when weighing credibility of critique | https://twitter.com/nephondemand/status/834238085986734082
- Errant Nephron @Errantnephron: this paper is worth a read and is #openaccess | https://twitter.com/Errantnephron/status/834320809242087424
- Ultimate Librarian @ultimatelibrarn: Fascinating. B/c I RT’d https://twitter.com/nephondemand/status/828236235365814272, I’m listed in this paper. #medlibs/#GCchat - thoughts on CME for Twitter journal clubs? | https://twitter.com/ultimatelibrarn/status/834279452028915712
- Willem Kolff @KidneyBeast: Impressive work. Shows how little value publishers add. They are in position to be disrupted. | https://twitter.com/KidneyBeast/status/834461886120939523
- Sylvi_ _enj_min @Gogmum: include how to #accredit your #twitter #journal #club | https://twitter.com/Gogmum/status/837026141466726413
- Katy Turner @katymeturner: thanks that’s really helpful! | https://twitter.com/katymeturner/status/847477183203295233
- Matt Graham-Brown @DrMattGB: Peer review is essential to maintaining research quality and integrity. Simply self-publishing circumvents this process | https://twitter.com/DrMattGB/status/902067552083472385 | AUTHOR RESPONSE: Self-publish doesn’t eliminate peer-review. p 4-6 of http://goo.gl/HU4WCP. Reviewers come from #SoMe community & write as much as they want | https://twitter.com/nephondemand/status/902087367787503617
- Kenar Jhaveri @kdjhaveri: How can one reach the readers you want with self publishing ? most nephrologist get @CJASN @AJKDonline and or @ISNkidneycare @NDTsocial | https://twitter.com/kdjhaveri/status/902016265119830018 | AUTHOR RESPONSE: See pg 7 of http://goo.gl/HU4WCP. Effective #SoMe use resulted in a reach of 40,221 readers in 15 days. That sufficiently counterbalances journal fees | https://twitter.com/nephondemand/status/902088596684984320

## AUDIENCE REACH

*Higher resolution image is found at the end of this document. Names in green denote individuals in Nephrology; black outside of Nephrology. Numbers in red indicate terminal audience reached*.

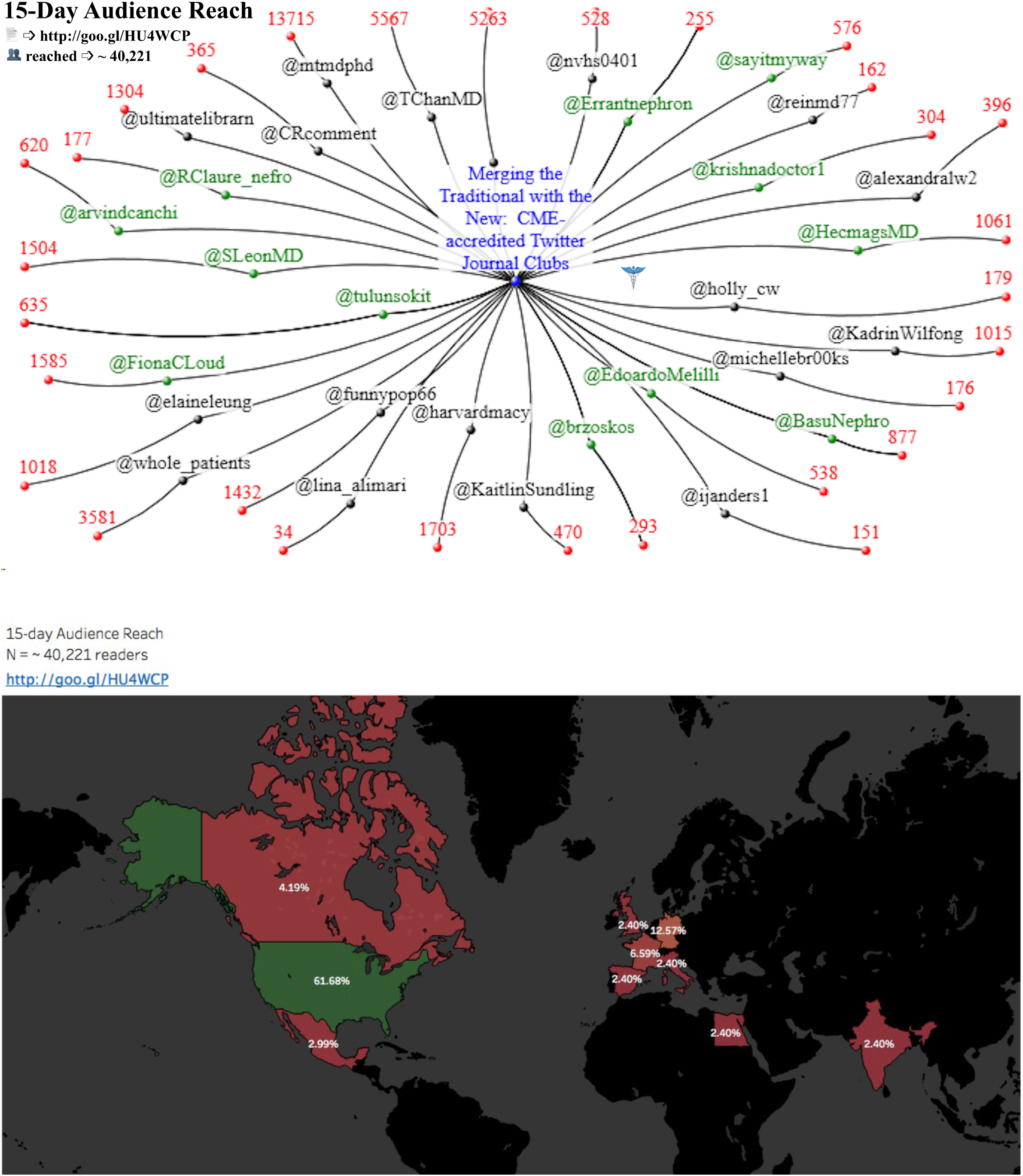

## TABLES

*Higher resolution images are found at the end of this document*.

## FIGURES

*Higher resolution images are found at the end of this document*.

## REFERENCES

1. Carpenter JP, Krutka DG. Chat it Up: Everything You Ever Wanted to Know about Twitter Chats but Were Afraid to Ask. Learning & Leading with Technology. 2014; 10–15.

2. Chan TM, Thoma B, Radecki R, et al. Ten Steps for Setting Up an Online Journal Club. Journal of Continuing Education in the Health Professions. 2015; 35:148–154.

3. Carpenter JP, Krutka DG. How and Why Educators Use Twitter: A Survey of the Field. Journal of Research on Technology in Education. 2014; 46:414–434.

4. Beckingham S, Nerantzi C, Reed P, et al. Speedy professional conversations around learning and teaching in higher education via the brand new tweetchat #LTHEchat. Sheffield Hallam University Research Archive. 2014. Available at: http://shura.shu.ac.uk/9337/. Accessed 26 December 2016.

5. Wood W. Online chats allow patients, physicians and advocates to share insights, experiences. HemeOnc Today. 2013; 32.

6. Micieli M, Frank JR, Jalali A. A Medical Educator’s Guide to #MedEd. Academic Medicine. 2015; 90:1176.

7. Roberts MJ, Perera M, Lawrentschuk N, et al. Globalization of Continuing Professional Development by Journal Clubs via Microblogging: A Systematic Review. Journal of Medical Internet Research. 2015; 17:e103.

8. Hillman T, Sherbino J. Social media in medical education: a new pedagogical paradigm? Postgraduate Medical Journal. 2015; 91:544–545.

9. Mehta N, Flickinger T. The Times They Are A-Changin’: Academia, Social Media and the JGIM Twitter Journal Club. J Gen Intern Med. 2014; 29:1317–1318.

10. Dalton M. “What Would I Tweet?”: Exploring New Professionals’ Attitudes Towards Twitter as a Tool for Professional Development. Journal of Library Innovation. 2013; 4:101–110

11. Perales MA, Drake EK, Pemmaraju N, et al. Social Media and the Adolescent and Young Adult (AYA) Patient with Cancer. Curr Hematol Malig Rep. 2016; 1–7.

12. Widmer RJ, Engler NB, Geske JB, et al. An Academic Healthcare Twitter Account: The Mayo Clinic Experience. Cyberpsychology, Behavior, and Social Networking. 2016; 19:360–366.

13. Leung EYL, Tirlapur SA, Siassakos D, et al. #BlueJC: BJOG and Katherine Twining Network collaborate to facilitate post-publication peer review and enhance research literacy via a Twitter journal club. BJOG. 2013; 120:657–660.

14. Lin M. Sherbino J. Creating a Virtual Journal Club: A Community of Practice Using Multiple Social Media Strategies. Journal of Graduate Medical Education. 2015; 481.

15. Ranschaert ER, van Ooijen Pma, Lee S, et al. Social media for radiologists: an introduction. Insights Imaging. 2015; 6:741–752.

16. Forgie SE, Duff JP, Ross S. Twelve tips for using Twitter as a learning tool in medical education. Medical Teacher. 2013; 35: 8–14.

17. Rogers S. What fuels a tweet’s engagement? Twitter Media Blog. Published March 10, 2014. Available at: https://blog.twitter.com/2014/what-fuels-a-tweets-engagement. Accessed 22 December 2016.

18. Gagnon K. Using Twitter in Health Professional Education. Journal of Allied Health. 2015; 44(1):25–33.

19. Reames BN, Sheetz KH, Engelsbe MJ, et al. Evaluating the Use of Twitter to Enhance the Educational Experience of a Medical School Surgery Clerkship. Journal of Surgical Education. 2015; 73:73–78.

20. Djuricich AM. Social Media, Evidence-Based Tweeting, and JCEHP. Journal of Continuing Education in the Health Professions. 2014; 34(4):202–204.

